# Null effect of circulating sphingomyelins on risk for breast cancer: a Mendelian randomization report using Breast Cancer Association Consortium (BCAC) data

**DOI:** 10.1101/19013540

**Authors:** Charleen D. Adams

## Abstract

**Background:** Changes in cellular metabolism are a hallmark of cancer and are linked with sphingolipid synthesis. Due to immense interest in how sphingolipids influence chemoresistance, more is known about the impact of sphingolipids during cancer treatment and progression than about the potential role of sphingolipids in the induction of tumors in humans.

**Methods:** Because estrogen triggers sphingolipid signaling cascades, the causal role of circulating levels of sphingomyelin (a type of sphingolipid) on breast cancer was investigated with a well-powered Mendelian randomization design.

**Results:** The results reveal a null effect (OR = 0.94; 95% CI = 0.85, 1.05; *P* = 0.30).

**Conclusion:** Despite the role sphingomyelins play during chemoresistance and cancer progression, circulating sphingomyelins do not appear to initiate or protect from breast cancer.

**Impact:** This finding comprises the first causal report in humans that sphingomyelins on breast cancer initiation is null. Future investigations of risk in other cancer types are needed to further explore the potential role of sphingolipid biology in cancer etiology.

---

Changes in cellular metabolism are a hallmark of cancer^1^. Sphingolipids can control the rate of cell proliferation during malignant transformation and affect chemoresistance^2^. Sphingomyelin is a type of sphingolipid, a class of lipids containing sphingoid bases (Fig. 1). As a response to cellular stress, sphingolipids mediate apoptosis and autophagy, through the synthesis and/or accumulation of ceramide. Ceramide can be hydrolyzed from sphingomyelin^3^. Due to immense interest in how sphingolipids influence chemoresistance, much is known about the impact of sphingolipids on cancer treatment and little is known about role sphingolipids in the induction of tumors in humans.

**Fig 1.**
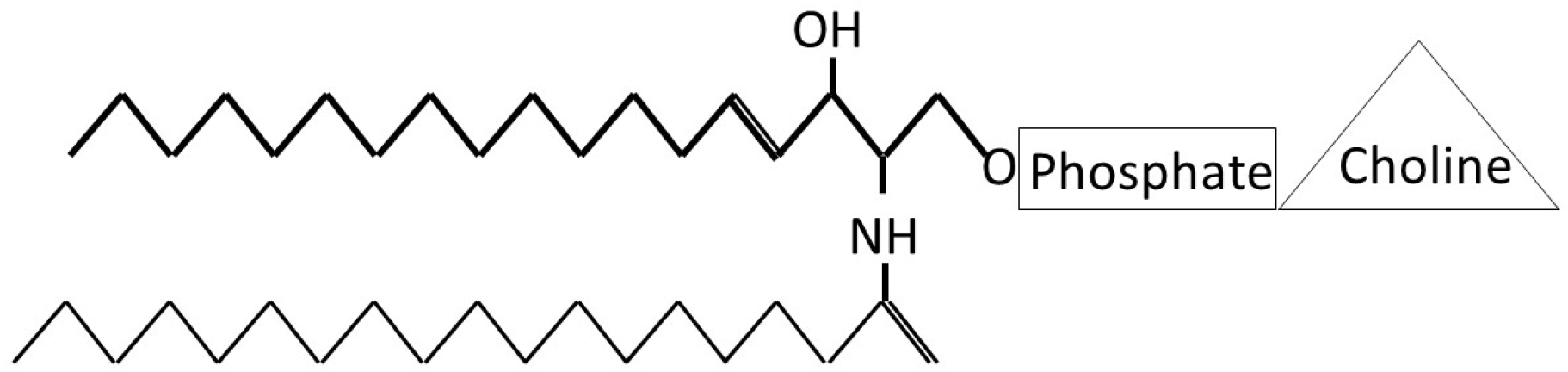
Cartoon of a sphingomyelin. The bolder print indicates the lipidic sphingoid base that is carrying a saturated fatty acid amine bonded to an amino group at the C2 position. Attached to the polar head is a phosphocholine. Figure adapted from Holthuis *et al*. (2001)^4^.

Estrogen triggers sphingolipid signaling cascades^2^. Due to this, it was hypothesized that circulating sphingomyelins might be involved in acquisition of breast cancer. The causal impact of circulating levels of sphingomyelins on risk for breast cancer was appraised with Mendelian randomization (MR).

## Materials and Methods

### Conceptual framework

MR is an instrumental variables technique; i.e., genetic variants (typically single-nucleotide polymorphisms, SNPs) strongly associated with traits are used in statistical models instead of the traits themselves. This avoids most environmental sources of confounding and averts reverse causation, which preclude causal inference in observational studies. Two-sample MR is an adaptation of the procedure that uses summary statistics from two genome-wide association (GWA) studies^5^.

### Mendelian randomization assumptions

MR depends on the validity of three assumptions: (i) the SNPs acting as the instrumental variables must be strongly associated with the exposure; (ii) the instrumental variables must be independent of confounders of the exposure and the outcome; and (iii) the instrumental variables must be associated with the outcome only through the exposure.

### Data sources

#### Step 1

Kettunen *et al*. (2016) performed a genome-wide association (GWA) study of 123 circulating metabolites—including sphingomyelins—in European participants (n=13,476 for sphingomyelins)^6^. From this, independent (those not in linkage disequilibrium; R^2^ < 0.01) SNPs associated at genome-wide significance (*P* < 5 × 10^−8^) with a standard-deviation (SD) increase in circulating sphingomyelins were identified. The Kettunen GWA is available through MR-Base (http://www.mrbase.org/)^5^.

#### Step 2

A publicly available GWA study of breast cancer performed by the Breast Cancer Association Consortium (BCAC) on 122,977 breast cancer cases and 105,974 controls of European ancestry was chosen as the outcome GWA for breast cancer^7^ (http://bcac.ccge.medschl.cam.ac.uk/).

### Statistical approach

A seven-SNP instrument for circulating sphingomyelins was constructed from the SNPs strongly associated with circulating sphingomyelin levels. Estimates of the proportion of variance in circulating sphingomyelins explained by the genetic instruments (R^2^) and the strength of the association between the genetic instruments and breast cancer (*F*-statistic) were generated (conventionally *F*-statistics <10 are weak). The instrument for sphingomyelins has an R^2^ = 0.032 and the *F*-statistic = 1089. The study was powered using the mRnd MR power calculator (available at http://cnsgenomics.com/shiny/mRnd/). It had >90% power to detect an OR of 0.90.

The log-odds for breast cancer per SD increase in circulating sphingomyelins was calculated, using the inverse-variance weighted (IVW) MR method. The “TwoSampleMR” package^5^ was used for the MR analysis.

All described analyses were performed in R version 3.5.2.

### Sensitivity analyses

Several sensitivity estimators can be used appraise pleiotropic bias. Three were chosen to complement the primary IVW causal tests: MR Egger regression, weighted median, and weighted mode estimations. In addition to these sensitivity estimators, a test for heterogeneity was performed, since variability in the causal estimates between SNPs can indicate pleiotropy. The test for heterogeneity was performed using Cochrane’s *Q*-statistic.

## Results

There was a null effect for circulating sphingomyelins on breast cancer (OR = 0.94; 95% CI = 0.85, 1.05; *P* = 0.30). The sensitivity estimators had effect estimates in the same direction and were of comparable magnitude to the IVW estimate, indicating no evidence for substantial bias due to unwanted pleiotropy. There was no evidence for heterogeneity in the estimates (Table 1). The MR-Egger intercept test, which provides an assessment of potential directional pleiotropy in the IVW was null. A null effect indicates a lack of evidence for pleiotropy (Estimate = 1.01; 95% CI = 0.97, 1.04; *P* =0.55).

**Table 1.** Causal estimates for the association of circulating sphingomyelin levels on risk breast cancer.

| Method | OR | Lower 95% CI | Upper 95% CI | P-value | Q-statistic | Q-diff | Q P-value |
| --- | --- | --- | --- | --- | --- | --- | --- |
| IVW | 0.94 | 0.85 | 1.05 | 0.30 | 8 | 6 | 0.27 |
| MR Egger* | 0.88 | 0.68 | 1.13 | 0.36 | 7 | 5 | 0.22 |
| Weighted median* | 0.92 | 0.81 | 1.04 | 0.19 | NA | NA | NA |
| Weighted mode* | 0.91 | 0.78 | 1.06 | 0.26 | NA | NA | NA |

IVW=inverse-weighted variance. \*Denotes a sensitivity estimator.

## Discussion

This is the first causal report in humans that sphingomyelins on breast cancer initiation is null. The null effect might reflect the complex interplay of pro-apoptotic and pro-growth ceramides^8^, perhaps with greater upregulation of the pro-apoptotic pathways, which may be different for different tissues. Future investigations of risk in other cancer types are needed to further explore the potential role of sphingolipid biology in cancer etiology.

## Data Availability

Data are publicly available via MR-Base and BCAC.

http://www.mrbase.org/

http://bcac.ccge.medschl.cam.ac.uk/

## Acknowledgements

The breast cancer genome-wide association analyses were supported by the Government of Canada through Genome Canada and the Canadian Institutes of Health Research, the ‘Ministère de l’Économie, de la Science et de l’Innovation du Québec’ through Genome Québec and grant PSR-SIIRI-701, The National Institutes of Health (U19 CA148065, X01HG007492), Cancer Research UK (C1287/A10118, C1287/A16563, C1287/A10710) and The European Union (HEALTH-F2-2009-223175 and H2020 633784 and 634935). All studies and funders are listed in Michailidou et al (2017).

## Notes

### Competing Interest Statement

The authors have declared no competing interest.

### Funding Statement

No funding received.

